# Factors associated with variation in single-dose albendazole pharmacokinetics: A systematic review and modelling analysis

**DOI:** 10.1101/2022.05.18.22275185

**Authors:** Charles Whittaker, Cédric B. Chesnais, Sébastien D.S. Pion, Joseph Kamgno, Martin Walker, Maria-Gloria Basáñez, Michel Boussinesq

## Abstract

**Background:** Albendazole is an orally administered anti-parasitic medication with widespread usage in a variety of both programmatic and clinical contexts. Previous work has shown the drug to be characterised by significant inter-individual pharmacokinetic variation. This variation is thought to have important consequences for treatment success, but current understanding of the factors associated with this variation remains incomplete.

**Methodology/Principal Findings:** We carried out a systematic review to identify references containing temporally disaggregated data on the blood concentration of albendazole and/or (its pharmacologically-active metabolite) albendazole sulfoxide following a single oral dose. These data were then integrated into a mathematical modelling framework to infer key pharmacokinetic parameters and relate them to characteristics of the populations being treated. These characteristics included age, weight, sex, dosage, infection status, and whether patients had received a fatty meal prior to treatment or other drugs alongside albendazole. Our results highlight a number of factors systematically associated with albendazole pharmacokinetic variation including age, existing parasitic infection and receipt of a fatty meal. These factors impact different aspects of the drug’s pharmacokinetic profile. Whilst age is significantly associated with albendazole sulfoxide half-life, receipt of a fatty meal prior to treatment was associated with increased albendazole bioavailability (and by extension, peak blood concentration and total drug exposure following the dose). Parasitic infection (particularly echinococcosis and neurocysticercosis) was associated with altered pharmacokinetic parameters, with infected populations displaying distinct characteristics to uninfected ones.

**Conclusions/Significance:** These results highlight the extensive inter-individual variation that characterises albendazole pharmacokinetics and provides insight into some of the factors associated with this variation.

**Author Summary:** Albendazole is a broad-spectrum anti-parasitic medication widely used in the treatment of a variety of parasitic worm infections. Previous studies have demonstrated significant variation in the pharmacokinetics of albendazole (and its pharmacologically active metabolite albendazole sulfoxide), leading to substantial inter-individual variability in blood plasma concentrations following a dose of the drug being given. This variation is thought to have important consequences for treatment success but our understanding of the factors driving this variation remain incomplete. In this study, we carried out a systematic review to identify references with data on albendazole and albendazole sulfoxide concentrations in the blood following a single oral dose. We then fitted a mathematical model of albendazole pharmacokinetics to these data to infer key pharmacokinetic parameters and relate them to characteristics of the populations being treated. We found that receipt of a fatty meal prior to treatment was associated with increased albendazole bioavailability, that the half-life of albendazole sulfoxide in the blood varied significantly with age, and that both echinococcosis and neurocysticercosis were associated with altered pharmacokinetic profiles compared to healthy individuals. Our work provides insight into some of the factors systematically associated with variation in albendazole pharmacokinetics.

## Introduction

Albendazole is a broad-spectrum medication used widely in the treatment of a variety of parasitic worm infections. This includes usage in a clinical context, where multiple-dose regimens are used to treat infections with the larval stages of *Taenia solium* ((neuro-)cysticercosis) [1] or of *Echinococcus* spp. (principally cystic and alveolar echinococcosis due to, respectively, *E. granulosus* and *E. multilocularis*) [2]. It has also been used extensively in neglected tropical disease (NTD) programmatic contexts, for which albendazole is being/has been delivered (as a single-dose treatment) to communities in mass drug administration (MDA) campaigns against soil-transmitted helminthiases [3] (STHs, due to *Ascaris lumbricoides, Trichuris trichiura, Necator americanus* and/or *Ancylostoma duodenale*), and delivered alone [4] or alongside ivermectin and/or diethylcarbamazine [5,6]) against lymphatic filariasis [7] (LF, due to *Wuchereria bancrofti* or *Brugia malayi*). In addition, albendazole has also been offered to individuals with loiasis [8,9], frequently those whose *Loa loa* microfilarial densities are high enough to preclude safe treatment with microfilaricidal anthelmintics [10,11] (such as diethylcarbamazine or ivermectin [12]).

Whilst the therapeutic efficacy of albendazole has been established for a wide array of helminth parasites, the drug’s pharmacokinetics (and those of its pharmacologically-active metabolite, albendazole sulfoxide) are characterised by extensive inter- and intra-individual variation. This variation has been consistently observed across a wide range of studies (see [13] for a review and its implications for treatment), and is typically attributed to the drug’s limited solubility in the gastrointestinal tract and extensive first-pass metabolism by the liver (responsible for rapid conversion of albendazole to albendazole sulfoxide). This variation is thought to contribute to the failure of cure in some treated patients – whilst some require only one course of treatment, others require multiple rounds and, in a limited number of instances, treatment failure has been observed [13–15]. This variation in outcomes observed in clinical settings has also been seen in field studies, where highly variable impact of the drug on STH infections have been observed depending on the setting [16]. For example, the average reduction in faecal egg reduction rates for treated hookworm infections varied from 53% to 95% across different communities in Ghana [17] (though note that such observations might also be driven by variation between settings in the prevalence of different STH species which are thought to show variable responses to albendazole treatment [18]).

A number of factors are deemed to underlie this variation in pharmacokinetic dynamics. Several studies have examined the influence of different drivers, including sex [19], co-administered drugs [20,21], delivery of albendazole alongside a fatty meal [22,23] and infection status [24,25] on the pharmacokinetic profile of albendazole and/or albendazole sulfoxide. However, these studies typically only analyse a single factor, and therefore a systematic understanding of the comparative impact of different factors on albendazole’s pharmacokinetics remains outstanding. Given albendazole’s widespread usage in NTD programmatic contexts, insight into mechanisms that influence the pharmacokinetic profile of albendazole could have significant public health relevance.

Motivated by this, we conducted a systematic review of the literature to identify references containing temporally disaggregated information on albendazole and/or albendazole sulfoxide concentrations in the blood following treatment with a single oral dose (the typical regimen used in NTD programmatic contexts). To these data, we fitted a mathematical model of albendazole and albendazole sulfoxide’s dynamics in the blood following receipt of the dose that captures key phenomena associated with the drug’s metabolism. These include albendazole’s extensive first-pass metabolism [26] and its established low bioavailability [27]. We fitted this model to data collated as part of the systematic review to infer key pharmacokinetic parameters, including albendazole bioavailability, albendazole sulfoxide half-life, peak albendazole concentration in the blood (*C*_*Max*_) and the total drug exposure across time (commonly described as the area under the curve or *AUC*). We then related these parameter estimates to characteristics of the patient populations being treated and the treatment regimen received.

## Methods

### Systematic review of albendazole pharmacokinetic literature

Web of Science and PubMed databases were searched on 4^th^ July 2019 with no limitations on date range using the keywords “albendazole” AND (treatment* OR dose* OR pharma* OR “half-life” OR “half life”) in order to identify references containing temporally disaggregated data detailing the concentration of albendazole and/or albendazole sulfoxide in the blood following treatment with a single dose of the drug. A total of 5690 unique records were identified through this search process, with 206 records retained for full text evaluation following Title and Abstract screening **(Fig 1)**. Studies lacking the required information on blood concentration levels over time, that had been carried out *in vitro* or in non-human subjects, or were not in English were subsequently excluded. Following this, a total of 32 references were included, yielding 92 time-series describing the evolution of blood concentrations of albendazole and/or albendazole sulfoxide following treatment with a single dose. Fifteen time-series contained information on both albendazole and albendazole sulfoxide levels; and 77 contained information on albendazole sulfoxide levels only. Eighty-four of these time-series were studies in which only blood drug concentrations following a single oral dose of albendazole were presented; and eight were studies in which measured blood drug concentrations were presented that spanned a course of multiple doses, but that contained information on drug blood concentration levels immediately following the first dose. For these eight time-series, we extracted data following receipt of the first dose up until receipt of the second dose. For each time-series, we also extracted the data describing evolution of albendazole/albendazole sulfoxide levels over time, as well as an array of metadata. These included characteristics of the treatment regimen (dose, fasting state, co-administered drugs), as well as information and metadata on the patients receiving treatment (sex, age, infection status and weight). In the majority of instances, presented data were reported for a population of patients rather than individuals. In these instances, population averages for factors such as age, weight, etc., were extracted. A full list of these references, as well as further information about each study and how the data were extracted are available in ***Supplementary S1 File, S1 Text: Data Extraction, Collation and Initial Processing***.

**Figure 1:**
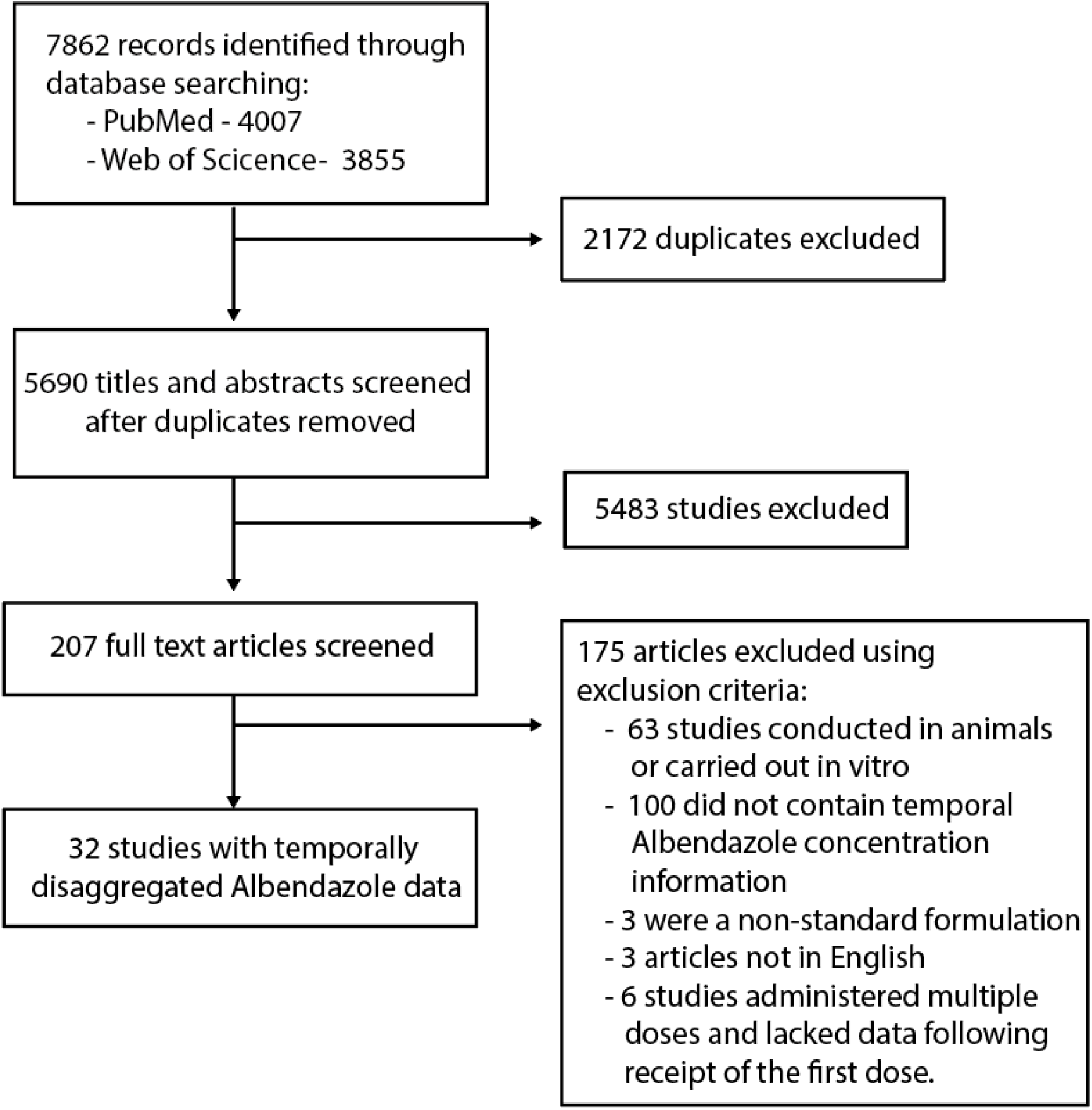
PRISMA diagram illustrating the systematic review workflow. Web of Science and PubMed were searched on 4^th^ July 2019 using the keywords albendazole AND (treatment* OR dose* OR pharma* OR “half-life” OR “half life”). This produced a total of 5690 results after duplicate removal, of which 206 were retained for full text screening. 175 of the retained articles were subsequently excluded based on pre-defined exclusion criteria, yielding 32 studies containing temporally disaggregated data on albendazole blood concentrations following treatment with a single dose; these 32 references contained a total of 92 time-series measuring albendazole and/or albendazole sulfoxide blood concentrations over time in different populations.

### Mathematical model construction and fitting

We developed a model describing the evolution of albendazole and albendazole sulfoxide concentrations in the blood following receipt of a single oral dose of the drug, based on a series of linked ordinary differential equations (ODEs) of albendazole and its metabolite albendazole sulfoxide **(Fig 2)**. The model incorporates a number of pharmacokinetic phenomena relevant to albendazole, including its well-established, limited bioavailability (thought to be a product of its poor solubility along the gastrointestinal tract [27]) and the extensive first-pass metabolism of albendazole to albendazole sulfoxide known to occur in the liver [26]. This model was fitted individually to each of the 92 collated time-series within a Bayesian framework, utilising an adaptive Metropolis-Hastings based Markov Chain Monte Carlo (MCMC) sampling scheme for parameter inference. Uninformative priors were used for each of the parameters being subsequently related to the collected metadata. For each dataset, a total of 25,000 iterations were run, with the first 5,000 discarded as burn in, leaving 20,000 samples available for parameter inference – the median value based on these samples was then used as an input into the multiple linear regression (described below). Further information on the exact formulation of the model and the fitting process is available in ***Supplementary S1 File, S2 Text: Model Construction, Fitting and Inference***.

**Figure 2:**
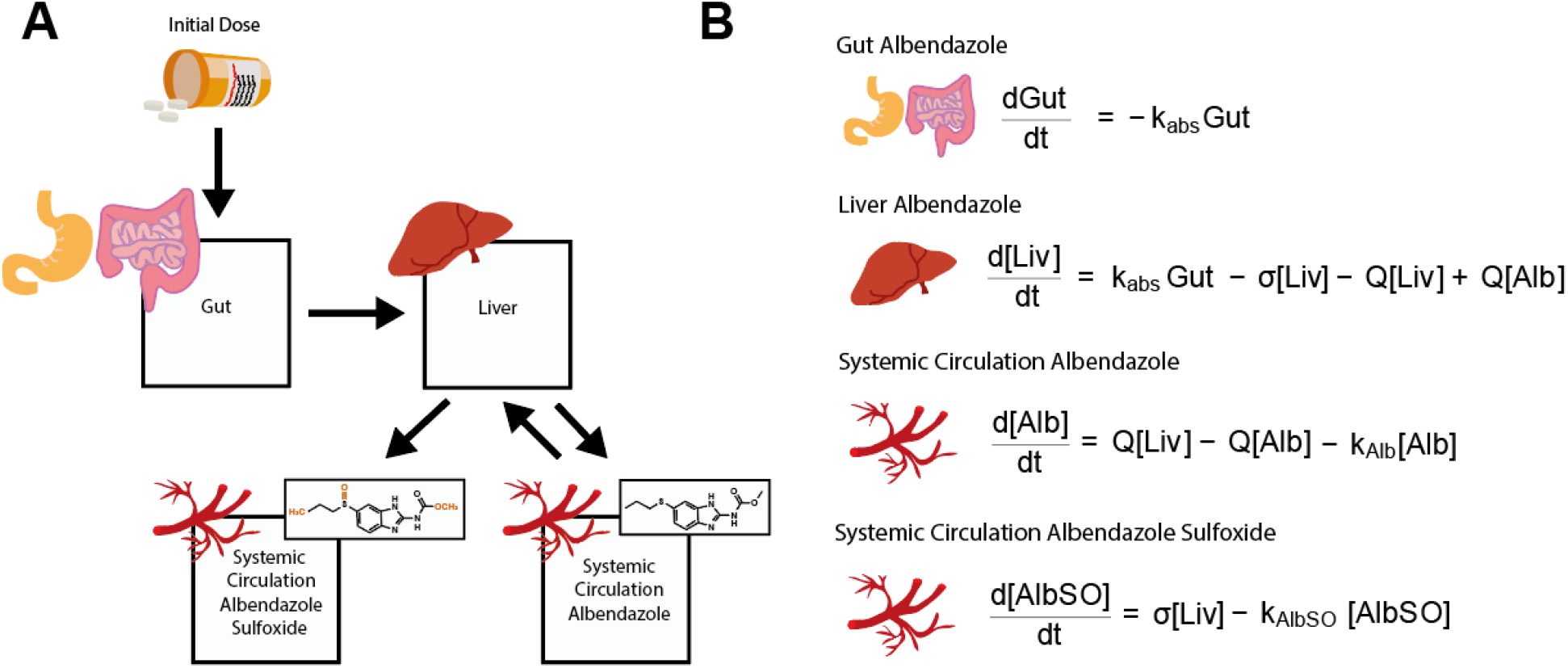
Schematic representation of the model describing albendazole and albendazole sulfoxide dynamics and pharmacokinetics. A compartmental model consisting of a series of linked ordinary differential equations (ODEs) was developed to simulate the pharmacokinetics of albendazole and its pharmacodynamically-active metabolite, albendazole sulfoxide, in the blood following a single oral dose. **(A)** Schematic illustrating the model structure and the way in which the different compartments are linked. **(B)** Overview of the ODEs governing the model, representing the amount of albendazole in the gut and concentration of the drug (and its metabolite albendazole sulfoxide) in the liver and systemic circulation. Note that the received dose is further scaled by a bioavailablity parameter not shown here – see ***Supplementary Information* Section “Mathematical Model of Albendazole and Albendazole Sulfoxide Dynamics”** for further details and full description of the model equations.

### Regression linking pharmacokinetic properties to patient characteristics

From the 92 fitted time-series, we extracted estimates of key pharmacokinetic parameters and regressed them onto the collected metadata (describing aspects of the patient population and treatment regimen received) to assess the influence of various factors on variation in albendazole and albendazole sulfoxide’s pharmacokinetics. The pharmacokinetic parameters were *k*_*AlbSO*_ (the half-life of albendazole sulfoxide), the *bioavailability* of albendazole (the proportion of administered albendazole absorbed from the gut into the blood), *C*_*Max*_ (the peak concentration of the drug in the blood) and *AUC* (“area under the curve”, reflecting the total exposure to the drug after administration of the dose, calculated over a time-period of 50 hours). *k*_*AlbSO*_ and *bioavailability* are model parameters directly estimated during the fitting process outlined above, and so for each time-series, the median parameter estimate from each time-series was used in the regression. For *C*_*Max*_ and *AUC*, in order to control for differences in dosages between studies (which would directly impact estimates of these two quantities), we used the fitted model (and median parameter estimates) for each time-series to simulate and generate a hypothetical pharmacokinetic curve assuming a standardised dose of 400 mg. We then calculated *C*_*Max*_ and *AUC* from this hypothetical pharmacokinetic curve to give estimates of the two parameters standardised by the dose received. We subsequently refer to these quantities as *C*_*Max*400_ and *AUC*_400_.

## Results

### Systematic review results and study characteristics

A total of 32 references containing 92 time-series detailing the concentration of albendazole and/or albendazole sulfoxide in the blood following treatment with a single dose of albendazole were identified. Forty-four time-series were data for a single individual and 58 time-series described average concentrations through time for a group of individuals (mean group size = 12.2, interquartile range = 6-14), with the data comprising a total number of 629 individuals who had received a single dose of albendazole. Of the 92 time-series identified, information on the sex of participants was available for 67 time-series (37 from male participants, 24 including a mixture of males and females, and 6 from female participants), with information on mean age and weight available for 79 and 69 time-series respectively. Sixteen time-series were from children under the age of 18. Information on whether treatment was taken with a fatty meal was available for 75 time-series (29 received a fatty meal, the remainder did not), whilst infection status was available for 91 time-series (48 were from uninfected patient populations, 16 were from individuals with neurocysticercosis, 14 from individuals with echinococcosis, 7 from individuals with onchocerciasis, 3 from individuals with lymphatic filariasis, 2 from individuals with giardiasis and 1 from an individual with hookworm infection). The median dose received was 400 mg (range 200 mg – 2205 mg). Co-administered drugs included ivermectin (n=7), diethylcarbamazine (DEC, n=7), praziquantel (n=4), ritonavir (n=2), dexamethasone (n=2), amoxicillin (n=1), gentamycin (n=1), metronidazole (n=1), ceftriaxone (n=1), levamisole (n=1) and oxantel pamoate (n=1). **Supplementary Table S1 in *Supplementary S1 File*** provides full details of each included study and time-series.

### Pharmacokinetic modelling of albendazole and albendazole sulfoxide dynamics

To each of these collated time-series, we fitted a model describing the dynamics of albendazole and albendazole sulfoxide concentrations in the blood following receipt of a single oral dose and before a second dose in the case of treatment regimen using multiple doses (see **Fig 2** for model structure and formulation). This model was fitted individually to each time-series within a Bayesian MCMC-based framework (see **Supplementary Fig S1** in ***Supplementary S1 File*** for individual model fitting results for each time-series). Our results highlighted significant variation in model estimates of key pharmacokinetic parameters including *k*_*AlbSO*_ (the half-life of albendazole sulfoxide), the bioavailability of albendazole (i.e. proportion of administered albendazole absorbed from the gut into the blood), *C*_*Max*400_ (peak modelled concentration of albendazole sulfoxide in the blood following receipt of a hypothetical 400 mg dose) and *AUC*_400_ (total modelled exposure to albendazole sulfoxide following receipt of a hypothetical 400 mg dose of albendazole). Stratifying the modelled pharmacokinetic profiles by various characteristics of the patient population suggested possible systematic pharmacokinetic differences associated with patient- and treatment regimen-related factors, although also extensive between-study variation in dynamics **(Fig 3)**.

**Figure 3:**
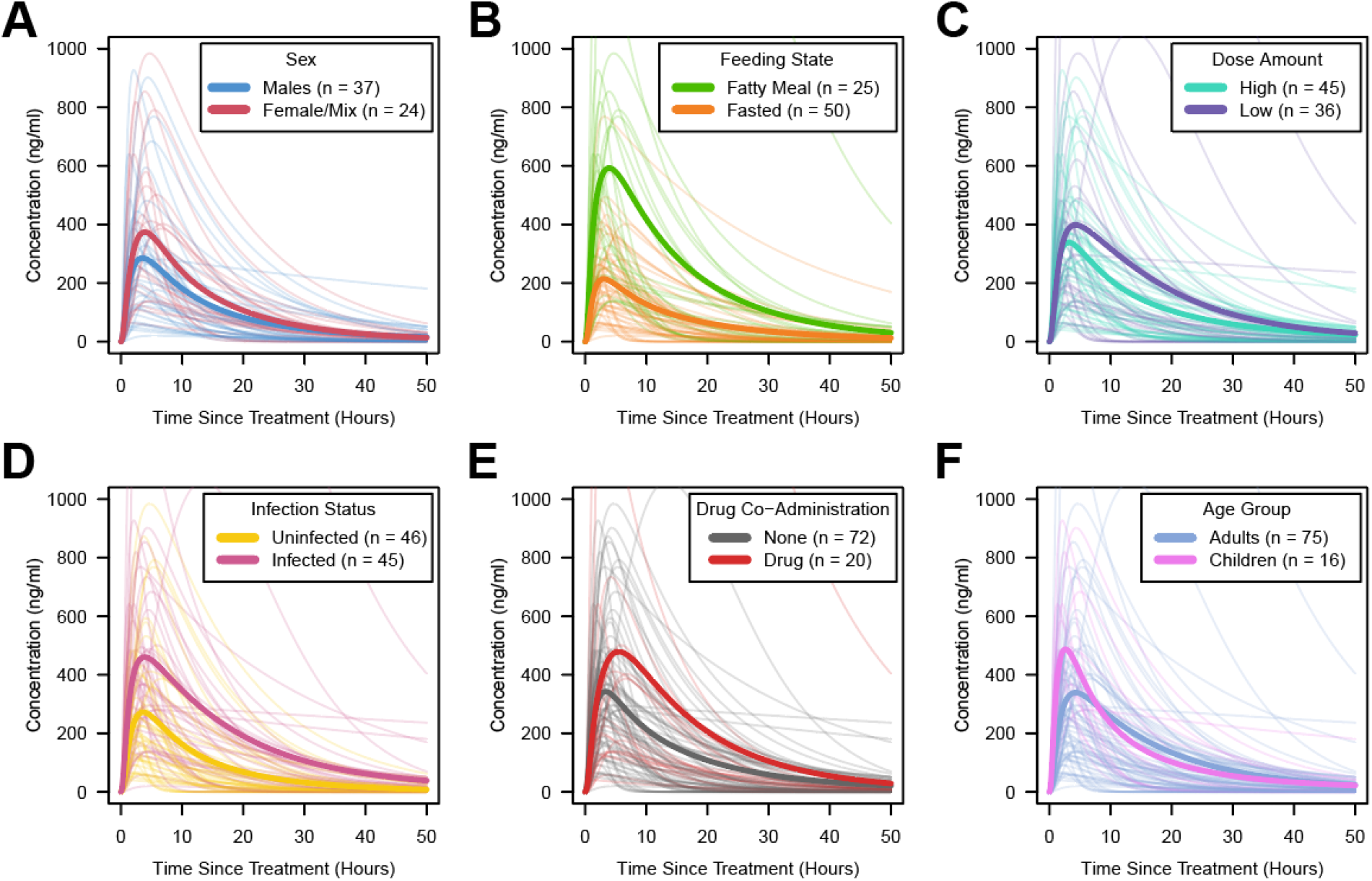
Albendazole sulfoxide pharmacokinetic variability, stratified by patient and dosage features. In all panels displayed above, each pale line represents the fitted model output for a single time-series, with the darker lines representing the average of the time-series for a given category. Factors explored were **(A)** Sex; **(B)** Feeding Status (according to whether groups had received the single dose of albendazole alongside a fatty meal or not); **(C)** Dose (with time-series crudely categorised into high/low strata based on whether the dose was higher than 400 mg); **(D)** Infection Status (defined as per the paper associated with each time-series as whether the patient population were receiving treatment for a known infection or not); **(E)** Co-Administered Drugs (i.e. whether albendazole was delivered alone or in tandem with other drugs); and **(F)** Age Group, defined based on whether the average age of the patients was under 18 years old (children) or ≥18 years (adults).

In order to explore these relationships more formally, we carried out a multiple linear regression analysis to assess which of the factors in **Fig 3** were statistically associated with differences in these pharmacokinetic parameters. The results of this regression are displayed in **Table 1**. Receipt of a fatty meal prior to treatment increased the bioavailability of albendazole by 38% on average (p<0.01) and resulted in a significantly higher peak blood concentration (*C*_*Max*400_ being 353 mg/ml higher or almost twice as high in individuals receiving a fatty meal on average, p<0.01). Receiving a fatty meal prior to treatment was also associated with a 1.53-fold higher overall *AUC*_400_ than in fasted individuals (p<0.01). Higher doses were associated with reduced albendazole bioavailability (with bioavailablity reducing approximately 1% for each 100 mg increase in dosage, p=0.03). We did not observe any significant differences in pharmacokinetic parameters that depended on sex.

**Table 1:** Regression outputs relating pharmacokinetic properties to characteristics. Inferred parameters from the fitted pharmacokinetic curves, specifically albendazole bioavailability, albendazole sulfoxide half-life, *AUC* and *C*_*Max*_ were regressed onto various patient population demographic and treatment metadata.

| | Bioavailability | AlbSO half-life | $AUC$ (for standardised 400 mg dose) | $C_{Max}$ (for standardised 400 mg dose) |
| --- | --- | --- | --- | --- |
| **Sex (Ref = male)** | p=0.25 | p=0.90 | p=0.44 | p=0.15 |
| **Fatty meal** | p=0.001 (+14%) | p=0.93 | p=0.01 (+6141) | p<0.001 (+330) |
| **Age group (Ref = adults)** | p=0.67 | p=0.19 | p=0.12 | p=0.95 |
| **Dose (mg)** | p=0.03 (-1.1% per 100 mg increase) | p=0.19 | p=0.30 | p=0.10 |
| **Parasitic infection (Ref = none)** | p=0.41 | p=0.10 | p=0.46 | p=0.95 |
| → **Onchocerciasis** | p=0.09 | p=0.48 | p=0.51 | p=0.08 |
| → **Echinococcosis** | p=0.04 (+14%) | p=0.15 | p<0.001 (+15604) | p<0.001 (+538) |
| → **Neurocysticercosis** | p=0.74 | p=0.05 (-1.5 hours) | p=0.32 | p=0.14 |
| **Co-administered drugs (Ref = none)** | p=0.09 | p=0.48 | p=0.58 | p=0.27 |

Parasitic infection was associated with significant differences in pharmacokinetic parameters compared to uninfected individuals. Whilst we did not detect any significant differences when considering infection status as a binary indicator (i.e. whether an individual had a parasitic infection or not), stratifying the infected population further by specific disease revealed significant associations between particular diseases. There was a significant association between neurocysticercosis and albendazole sulfoxide half-life (median 12 hours compared to 10.5 hours in uninfected individuals, p=0.05); and significant effects of echinococcosis on bioavailability (increased by 14% in infected compared to uninfected individuals, p<0.01) and *C*_*Max*400_ and *AUC*_400_ (increased by 2.7 and 4.0 fold respectively, p<0.01 in both instances). We did not observe any significant association between onchocerciasis and the considered pharmacokinetic parameters.

As a sensitivity analysis, we repeated the analyses described above controlling for the dose of albendazole received per kilogram of body weight (available only for a subset of the time-series due to a lack of complete information about participants’ weight), rather than the raw amount (in mg, not standardised by body weight) given to an individual. All significant associations described above were retained when conducting this subset sensitivity analysis (see **Supplementary S2 Table** in ***Supplementary File S1***). Additionally, we observed a difference between age groups in the modelled estimates of *k*_*AlbSO*_, with the median half-life of albendazole sulfoxide at 12.4 hours in adults compared to only 8.04 hours in children under the age of 18 years (p<0.01).

We did not detect a significant effect of co-administered drugs on albendazole’s pharmacokinetics, though it is important to note that the heterogeneous array of drugs co-administered across the collated dataset, and the comparative paucity of time-series featuring each of the drugs precluded a stratified analysis of each drug individually (as was possible with disease status). This lack of data necessitated combining them into the binary category or yes/no co-administration. The corollary of this is that these analyses were not powered to reliably detect drug-drug interactions with albendazole (which are well documented in the literature).

## Discussion

Despite widespread usage, significant uncertainty surrounds the factors underlying variation in the pharmacokinetics of albendazole. Whilst other studies have previously examined these factors individually (e.g. [23,28–31] amongst others), a systematic analysis of different factors together remained outstanding. Integrating the results of a systematic review of the literature with a mathematical model of albendazole/albendazole sulfoxide dynamics, our work highlights the extensive inter-individual pharmacokinetic variation in albendazole’s dynamics, and the impact that a number of different factors has in shaping the pharmacokinetic profile of the drug (and its metabolite) in the blood following receipt of a single oral dose. Importantly, our results suggest that these different factors influence different pharmacokinetic parameters, and hence alter different aspects of the pharmacokinetic profile of the drug in the blood.

In-keeping with previous work [22,23,32–34], consumption of a fatty meal prior to receiving the dose was associated with increased bioavailability of albendazole, increasing the amount absorbed into the body (and concomitantly elevating the *AUC* and *C*_*Max*_ values achieved), a phenomenon thought to be attributed to changes in the drug’s solubility (previously shown to be the rate-limiting step in albendazole’s bioavailablity and absorption [27]) when delivered alongside a fatty meal [34]. Whilst prior results from the literature have suggested (modest) differences between men and women in albendazole’s pharmacokinetics (specifically with regards to the *AUC* and *C*_*Max*_ [19]), we did not observe any statistically significant differences here. However, important caveats to our results are that the lack of individual data in many cases precluded examination of men and women separately. Therefore, we constructed a crude proxy for comparison (between men and groups in which the population comprised mixtures of men and women), which may not have been powered to detect the (minor) differences previously reported [19]. We observed a small but significant effect of the dose of albendazole received on the bioavailability of the drug, with bioavailability decreasing as the dose increased. This is consistent with the well-described induction by albendazole of enzymes responsible for its own metabolism [35]. Together, our results also suggest that different factors impact different pharmacokinetic properties of albendazole and albendazole sulfoxide. For example, whilst receipt of a fatty meal was associated with increases to bioavailability, *AUC* and *C*_*Max*_, age was significantly associated with albendazole sulfoxide half-life (when controlling for dosage per kilogram of body weight).

Perhaps most interestingly, our analyses suggested significant effects of parasitic infection on albendazole pharmacokinetics, with the exact impact dependent on the infection being considered. Previous work in sheep has highlighted that gastrointestinal nematode infection can influence the kinetics of albendazole and albendazole sulfoxide, leading to increased *AUC* values [36]. Other work has noted increased rates of transformation from albendazole to albendazole sulfoxide in infected compared to uninfected sheep [37], though this has been inconsistently observed and the exact influence (or lack thereof) likely depends on the particular infecting parasite [38]. Work in humans has suggested that the precise impact of infection depends on the interaction between the drug (particularly its absorption and elimination) and the infecting parasite’s impact on the host. For example, whilst recent work comparing the pharmacokinetics of albendazole in uninfected and *Wuchereria bancrofti*-infected adults showed no differences [24], previous work exploring albendazole kinetics in patients with echinococcosis demonstrated delayed absorption and impaired elimination of the drug (with this latter effect contributing to increases in the *AUC* of albendazole sulfoxide, particularly in patients with hepatic obstruction due to the disease) [39]. In-keeping with these results, we observed a significant effect of echinococcosis on albendazole’s pharmacokinetic parameters, with infection associated with increases to the bioavailability, C_Max_ and *AUC* of the drug. By contrast, we observed no significant effect of onchocerciasis on albendazole’s pharmacokinetics. For neurocysticercosis, we observed alterations to the apparent half-life of albendazole sulfoxide. However, these results should be interpreted with caution. Sample sizes for each of the individual infections were small; for example, the largest was for echinococcosis with 14 time-series drawn from a total of four studies. The estimates presented here are therefore uncertain, and it is possible that study-specific variation not accounted for might explain the observed results. Relatedly, whilst we attempted to control for co-administered drugs, our ability to do this was limited (see below). It is, therefore, possible that the results presented here might be confounded by the receipt of treatment for an infection that is not described in the reference.

There are a number of limitations to the analyses presented here. Firstly, and perhaps most notably, the available data in the literature were highly heterogeneous, involving a diversity of treatment regimens (i.e. other co-administered drugs) and patients (i.e. characteristics), with data available at different levels of aggregation (i.e. individual vs. average profiles). This constraint limits the statistical power of our analyses to characterise the effects of different individual drugs on albendazole’s pharmacokinetics. Whilst our binary indicator for co-administered drugs was not found to be significantly associated with any of the pharmacokinetic parameters explored here, numerous interactions between albendazole and other drugs such as cimetidine [20], azithromycin [40] and various anti-epileptic drugs [41] are well-documented in the literature. It is also important to note the overall paucity of available data upon which the inferences presented here are based. Whilst albendazole sulfoxide data were available for all 92 time-series considered here, data on albendazole’s pharmacokinetics was only available for 15 time-series; and in no cases was comparable information relevant to the drug’s metabolism (such as liver function) available.

In addition to these constraints posed by population-level data, the results presented here are limited in that they only describe the pharmacokinetic dynamics following treatment with a single dose of albendazole. This holds programmatic relevance given usage of albendazole in MDA programmes targeting STH [42] and/or LF [43] amongst others but other treatment regimen exist, most notably the use of albendazole in dedicated clinical settings to treat individuals for diseases such as cysticercosis and echinococcosis. These regimens typically utilise multiple doses delivered over consecutive days. Previous results have indicated that albendazole appears to induce its own metabolism through induction of key enzymes in the liver [25], and that multiple doses given over sequential days can lead to changes in pharmacokinetic properties over the course of multiple dose regimen; specifically, reductions in the maximum blood concentrations of albendazole sulfoxide reached following each dose [44]. However, the magnitude of this effect and the frequency of dosing required to elicit pharmacologically-relevant reductions in blood concentrations remain far from clear and have, to date, been addressed in only a limited number of studies. Exploration of this phenomenon and its consequences for anthelmintic treatment regimens using multiple doses of the drug would require both further clinical research and an extension of the mathematical model developed here, and likely represents an instructive avenue of future investigation. Similarly, extensions of the model to include infrequent but reported pharmacokinetic phenomena associated with albendazole treatment, such as biphasic pharmacokinetic profiles (thought possibly to be a product of inter-individual variation in frequency of gastric emptying affecting release of ingested albendazole into the gut, as well as other related characteristics [45]), would also likely provide new insight.

The availability of studies explicitly exploring the pharmacokinetics of drugs used to treat NTDs is limited [46]. Despite the limitations described above, our work begins to address this gap for albendazole. It suggests potential useful avenues for improvements to programmatic delivery of albendazole, and perhaps more importantly, highlights the existence of significant inter-individual variation in albendazole pharmacokinetics. Whilst we provide insight into some of the factors underlying this variation, further quantification and exploration will be essential. This is particularly crucial for understanding and interpreting the results of studies exploring programmatic usage of the drug to treat parasitic infections. Having an understanding of the degree of inter-individual variation will be vital for establishing whether observed sub-optimal responses to the drug are due to parasitic factors (e.g. possible resistance potentially developed through cumulative exposure to treatment over multiple rounds [47]) or, instead, simply reflect a high degree of variation between individuals in total exposure to the drug (and hence anti-parasitic effect) that follows ingestion of the same oral dose. Indeed, previous work has demonstrated a high degree of community-level variation in the impacts of ivermectin distribution campaigns to treat onchocerciasis in Ghana, even across communities characterised by similar histories of ivermectin exposure [48], highlighting the need to understand patterns of variation in drug responses (both at the individual and community level) if the underlying causes are to be accurately determined. Together, these results support and underscore recent calls highlighting the need for the collection, collation and analysis of individual participant data (IPD) to generate robust evidence on efficacy and safety of anti-parasitic treatment regimens [49]. Given the increasing frequency with which albendazole is being utilised as part of community-based MDA programmes aimed at controlling a wide array of parasitic infections and NTDs, such an understanding would hold important public health relevance.

## Data Availability

We attach the data in the supplementary information of this submission. And also, all the data, analyses, code etc can be found at https://github.com/cwhittaker1000/albendazole_pk

## Data and Code Availability

All data collated as part of this study, as well as the analytical code used to generate the analyses reported in the Main Text and Supplementary File can be found at the following link: https://github.com/cwhittaker1000/albendazole_pk.

## Acknowledgements

We deeply thank Dr Annette C. Kuesel for her hugely insightful and helpful comments on earlier versions of this manuscript, which have materially improved and contributed to the work now presented here.

## Authors Contributions

**Charlie Whittaker:** Conceptualization, Data Curation, Formal Analysis, Investigation, Methodology, Software, Visualization, Writing – Original Draft Preparation; **Cédric B. Chesnais:** Resources, Writing – Review & Editing; **Sébastien D.S. Pion:** Writing – Review & Editing; **Joseph Kamgno:** Writing – Review & Editing; **Martin Walker:** Resources, Writing – Review & Editing; **Maria-Gloria Basáñez:** Conceptualization, Funding Acquisition, Project Administration, Resources, Supervision, Writing – Review & Editing; **Michel Boussinesq:** Conceptualization, Project Administration, Resources, Supervision, Writing – Review & Editing.

## Supplementary S1 File: Additional Methods and Results

**S1 Text: Data Extraction, Collation and Initial Processing**

***Systematic Review References & Associated Metadata***

**S1 Table: Studies collated through the systematic review and their associated metadata**

**S2 Text: Model Construction, Fitting and Inference**

***Mathematical Model of Albendazole and Albendazole Sulfoxide Dynamics***

***Model Fitting and Inferential Framework***

***Pharmacokinetic Parameter Estimation and Multiple Linear Regression Modelling***

**S1 Figure: Results of model fitting and calibration to data collated through the systematic review**.

**S2 Table: Multiple linear regression results relating pharmacokinetic properties to study characteristics when controlling for dosage per kilogram of body weight instead of raw dosage amount in milligrams**

**Supplementary References**

