## Supplementary Information for "Factors associated with variation in single-dose albendazole pharmacokinetics: A systematic review and modelling analysis"

**Supplementary File S1: Additional Methods and Results**

**Factors associated with variation in single-dose albendazole pharmacokinetics: A systematic review and modelling analysis**

**Short title: Review and modelling of albendazole pharmacokinetics**

Charles Whittaker^1,§^**,** Cédric B. Chesnais^2^, Sébastien D.S. Pion^2^, Joseph Kamgno^3^, Martin Walker^4,5^, Maria-Gloria Basáñez^1,5^* & Michel Boussinesq^2^*

^1^ MRC Centre for Global Infectious Disease Analysis, Department of Infectious Disease Epidemiology, School of Public Health, Imperial College London, London, UK

^2^ Institut de Recherche pour le Développement (IRD), Montpellier, France

^3^ Centre for Research on Filariasis & other Tropical Diseases, and Faculty of Medicine and Biomedical Sciences, University of Yaoundé I, Yaoundé, Cameroon

^4^ Department of Pathobiology and Population Sciences, Royal Veterinary College, Hatfield, UK

^5^ London Centre for Neglected Tropical Disease Research, Department of Infectious Disease Epidemiology, School of Public Health, Imperial College London, London, UK

* Joint senior authors

**Contents:**

**S1 Text: Data Extraction, Collation and Initial Processing**

**S1 Table: Studies collated through the systematic review and their associated metadata**

**S2 Text: Model Construction, Fitting and Inference**

**S1 Figure: Results of model fitting and calibration to data collated through the systematic review**

**S2 Table: Multivariate linear regression results relating pharmacokinetic properties to study characteristics when controlling for dosage per kilogram of body weight instead of raw dosage amount in milligrams.**

**Supplementary References**

**Outline of Document**

In this **Supplementary S1 File** we describe the methods and data used to explore and analyse the drivers of variation in albendazole (and albendazole sulfoxide) pharmacokinetics. In **S1 Text: Data Extraction, Collation and Initial Processing**, we present further information on the systematic review conducted, including details of the collated references and information on the metadata (population characteristics, infection status, co-administration of other drugs etc.) available for each study. In **S2 Text: Model Construction, Fitting and Inference**, we detail the mathematical and statistical methodologies employed to process these extracted data, whose output forms the basis for the results presented in the Main Text. This includes further details on the pharmacokinetic model, the Bayesian fitting procedures and the multivariate linear regression analysis relating results from the fitting to study metadata. **S1 Table**, **S1 Figure** and **S2 Table** present additional results to support the work detailed in the Main Text. Finally, the **Supplementary References** provide details on the papers listed in **S1 Table** in addition to other references mentioned in this document.

**S1 Text: Data Extraction, Collation and Initial Processing**

***Systematic Review References & Associated Metadata***

We searched the Web of Science and PubMed databases on 4^th^ July 2019 with no date constraints using the keywords “albendazole” AND (treatment* OR dose* OR pharma* OR “half-life” OR “half life”) in order to identify references containing temporally disaggregated data detailing the concentration of albendazole and/or albendazole sulfoxide in the blood following treatment with a single dose of the drug. References were selected for Inclusion/Exclusion according to the following criteria:

**Inclusion Criteria:**

- Reference contains data from human subjects describing the concentration of albendazole and/or albendazole sulfoxide in the blood following receipt of a single, orally administered dose of albendazole.

**Exclusion Criteria:**

- The study was carried out in animals or *in vitro*, i.e. not in humans.
- The study administered multiple doses of albendazole and does not contain information on blood drug concentration following receipt of the very first dose.
- Reference does not contain temporally disaggregated information on albendazole/albendazole sulfoxide concentrations in the blood.
- The article is not in English.

A total of 7862 records were identified, with 2172 duplicates being excluded, leaving 5690 unique records being retained for title and abstract screening. Title and abstract screening excluded 5483 references, leaving a total of 207 articles for full-text screening. Studies lacking the required information on blood concentration levels over time, not in English, that utilised non-standard formulations of albendazole (e.g. oral suspension), or that had been carried out *in vitro* or in non-human subjects were subsequently excluded. A total of 32 references were subsequently retained and included for data extraction. For each reference, we extracted all relevant albendazole and albendazole sulfoxide concentration data over time that were available, yielding 92 time-series describing the evolution of blood concentrations of albendazole (n=15) and/or albendazole sulfoxide (n=92) in individuals or groups of individuals following treatment with a single dose. For each time-series, we also extracted relevant metadata and characteristics of the individual/group of individuals receiving treatment. These metadata were:

- **Sex:** The sex of the individual, or composition of sexes in the case of groups of individuals. This was subsequently converted into a categorical variable based on the collated responses, with levels “Males” (where the entire population sample consisted of male subjects), “Mixture” (where the population sample was a mixture of males and females) and “Unclear” (where sex of the individual/group was not provided), for use in the regression analyses.
- **Age:** The age of the individual, or in the case of groups of individuals, the mean age of the individuals. This was subsequently converted into a binary indicator according to whether the age of individuals was ≥18 (“Adults”) or < 18 years (“Children”), for use in the regression analyses.
- **Dose Amount:** Both the total dose amount (in mg) and the dose per kilogram of bodyweight. Where the latter was not directly provided but the weight of participants was provided, the dose per kilogram of bodyweight was calculated manually.
- **Feeding State:** Whether or not the individual or group of individuals had received a fatty/oily meal prior to receiving albendazole.
- **Co-Administered Drugs:** Details on whether or not the reference reported any drugs that either 1) were co-administered alongside albendazole or 2) which the individuals were receiving prior to receiving albendazole, and continued to take following receipt of the albendazole dose. This was subsequently converted into a binary indicator denoting whether or not any drugs were being taken alongside albendazole (“Yes”/“No”).
- **Infection Status:** Details on whether or not the reference reported that the individual or group of individuals receiving albendazole were doing so because they currently had a parasitic infection (as defined by the reference) and, if so, what parasite species they were infected with. As with co-administered drugs, this was also converted into a binary indicator denoting whether or not the individual or group of individuals had a reported parasitic infection.
- **Weight:** Where available, we also collated and extracted information on the weight of an individual, or the average weight of a group of individuals.

We extracted albendazole and albendazole sulfoxide blood concentration data at the individual level where possible, only extracting this information for groups of individuals where individual-disaggregated data were not available. Where data were presented at the individual level but only group-level characteristics (such as age, sex or weight) were present, we associated each individual-level time-series with the relevant group-level average characteristic. Where individuals had received multiple doses of albendazole but where there were pharmacokinetic data describing blood concentrations following the first dose, we extracted information on blood concentrations for all time-points up until receipt of the second dose.

**Supplementary Table S1: Studies collated through the systematic review and their associated metadata.** Further details and disaggregation of metadata by each specific time-series (rather than reference) is also available here: <https://github.com/cwhittaker1000/albendazole_pk>.

| **First Author [Ref]** | **Year** | **# Time-Series** | **Total Number of Individuals** | **Drug Blood Concentration Information** | **Dose (mg)** | **Dose (per kg)** | **Sex** | **Age**  **(years)** | **Fatty Meal?** | **Co-Drugs** | **Infection** |
| --- | --- | --- | --- | --- | --- | --- | --- | --- | --- | --- | --- |
| Awadzi [1] | 2003 | 2 | 28 | AlbSO Only | 400 | Various | All Males | Various (Adults) | None | Various (None; IVM, DEC & PZQ) | Onchocerciasis |
| Awadzi [2] | 2004 | 1 | 22 | AlbSO Only | 400 | 7.49 | All Males | 43 (Adults) | None | Levamisole | Onchocerciasis |
| Awadzi [3] | 1994 | 2 | 28 | AlbSO Only | 1200 | NA | All Males | NA (Adults) | Various | None | Onchocerciasis |
| Ceballos [4] | 2018 | 1 | 8 | Alb & AlbSO | 400 | 6.25 | Mixture | NA (Adults) | None | None | None |
| Chen [5] | 2004 | 1 | 20 | Alb & AlbSO | 400 | NA | All Males | NA (Adults) | NA | None | None |
| Chhonker [6] | 2018 | 1 | 7 | Alb & AlbSO | 400 | NA | Mixture | NA (Adults) | None | IVM | Mixture (None; Lymphatic filariasis) |
| Corti [7] | 2009 | 3 | 24 | Alb & AlbSO | 400 | 5.48 | All Males | 31 (Adults) | None | Various (None; Ritonavir) | None |
| Cotting [8] | 1990 | 3 | 3 | AlbSO Only | 200 | Various | All Females | Various (Adults) | Fatty Meal | Various (Amoxicillin & Gentamicin; Metronidazole and Ceftriaxone) | Echinococcosis |
| Delatour [9] | 1991 | 1 | 4 | AlbSO Only | 725 | 10 | All Males | Various (Adults) | None | None | None |
| Edi [10] | 2019 | 2 | 56 | Alb & AlbSO | 400 | NA | Mixture | Various (Adults) | NA | IVM & DEC | Various (None; Lymphatic filariasis) |
| Hoaksey [11] | 1991 | 2 | 32 | AlbSO Only | Various | Various | All Males | 37 (Adults) | Fatty Meal | None | Onchocerciasis |
| Jung [12] | 1992 | 8 | 8 | AlbSO Only | Various | 15 | Mixture | Various (Adults) | Fasted | None | Neurocysticercosis |
| Jung [13] | 1997 | 8 | 8 | AlbSO Only | Various | 15 | Mixture | Various (Children) | Fatty Meal | None | Neurocysticercosis |
| Kitzman [14] | 2002 | 1 | 1 | Alb & AlbSO | 400 | NA | NA | NA | NA | None | None |
| Lange [15] | 1988 | 2 | 12 | AlbSO Only | 400 | 5.65 | Mixture | 43.5 (Adults) | Various | None | None |
| Monteiro [16] | 2010 | 2 | 18 | AlbSO Only | 400 | 6.23 | Mixture | 26 (Adults) | None | Various (None; PZQ) | None |
| Marriner [17] | 1986 | 10 | 10 | AlbSO Only | 400 | 5.93 | NA | 27.5 (Adults) | Various | None | None |
| Mingjie [18] | 2002 | 1 | 7 | AlbSO Only | 830 | 12.5 | All Males | 29.3 (Adults) | NA | None | Echinococcosis |
| Mirfazaelian [19] | 2002 | 3 | 30 | AlbSO Only | Various | Various | Mixture | 32.5 (Adults) | None | None | None |
| Mirfazaelian [20] | 2003 | 2 | 12 | AlbSO Only | 800 | 11.81 | Mixture | 30 (Adults) | Fasted | None | None |
| Na-Bangchang [21] | 2006 | 2 | 46 | AlbSO Only | 400 | 7.53 | Mixture | 21 (Adults) | None | Various (IVM; IVM & PZQ) | None |
| Nagy [22] | 2002 | 4 | 24 | AlbSO Only | 690 | 10 | All Males | 20 (Adults) | Various | Various (None; Cimetidine) | None |
| Okelo [23] | 1993 | 5 | 5 | AlbSO Only | 250 | 9.33 | All Males | 9.5 (Children) | NA | None | Echinococcosis |
| Pengsaa [24] | 2004 | 2 | 20 | Alb & AlbSO | 400 | Various | Mixture | Various (Children) | Fatty Meal | Various (None; PZQ) | Giardiasis |
| Rathod [25] | 2016 | 1 | 51 | Alb & AlbSO | 400 | NA | NA | NA (Adults) | None | None | None |
| Rigter [26] | 2004 | 1 | 1 | AlbSO Only | 400 | 7.14 | NA | 29.5 (Adults) | None | None | None |
| Sarin [27] | 2004 | 1 | 10 | AlbSO Only | 600 | 10.03 | NA | 32.5 (Adults) | Fatty Meal | None | None |
| Schipper [28] | 2000 | 9 | 30 | AlbSO Only | Various | Various | All Males | 20 (Adults) | None | None | None |
| Schulz [29] | 2019 | 1 | 10 | Alb & AlbSO | 400 | NA | NA | 16.5 (Children) | None | Oxantel pamoate | Hookworm infection |
| Sergio- Mares [30] | 2005 | 2 | 32 | AlbSO Only | 800 | 12.72 | Mixture | 24.7 (Adults) | Various | None | None |
| Shenoy [31] | 2002 | 2 | 28 | Alb & AlbSO | 400 | Various | Mixture | 31.5 (Adults) | None | Various (None; DEC) | None |
| Thomsen [32] | 2016 | 2 | 24 | AlbSO Only | 400 | Various | Mixture | Various (Adults) | None | Various (None; IVM & DEC) | Lymphatic filariasis |

Alb: Albendazole; AlbSO: Albendazole sulphoxide; IVM: ivermectin; DEC: diethylcarbamazine; PZQ: praziquantel; NA: not available.

**Supplementary Text S2: Model Construction, Fitting and Inference**

***Mathematical Model of Albendazole and Albendazole Sulfoxide Dynamics***

A mathematical model describing the evolution of albendazole and albendazole sulfoxide concentrations in the blood following receipt of a single dose, based on series of linked ordinary differential equations (ODEs) was developed. The model included a number of pharmacokinetic dynamics features known to be relevant to albendazole, including its limited bioavailability (which is thought to be due to its poor solubility along the gastrointestinal tract) [33] as well as first-pass metabolism of albendazole to albendazole sulfoxide known to occur via the liver [34]. In brief, following administration of an oral dose of albendazole, we model the amount of drug in the gut, and its subsequent absorption into the body. We model the newly absorbed albendazole as passing directly through a liver compartment that converts some proportion of passaged albendazole into the metabolite albendazole sulfoxide via first-pass metabolism. Subsequent circulation and exchange of peripheral and hepatic blood leads to further conversion of albendazole into albendazole sulfoxide. Additionally, we model both albendazole and albendazole sulfoxide as being metabolised by enzymatic processes, leading to gradual removal over time. The model is specified mathematically as follows,

| $\frac{d{Alb}_{Gut}}{dt}= -k_{Abs}{Alb}_{Gut}$ | Eqn. [S1] |
| --- | --- |
| $\frac{d[{Alb}_{Liver}]}{dt}= k_{Abs}{Alb}_{Gut}- \sigma\left[ {Alb}_{Liver} \right]-Q\left[ {Alb}_{Liver} \right]+Q[{Alb}_{Periph}]$ | Eqn. [S2] |
| $\frac{d[{Alb}_{Periph}]}{dt}= Q\left[ {Alb}_{Liver} \right]-Q\left[ {Alb}_{Periph} \right]-k_{Alb}[{Alb}_{Periph}]$ | Eqn. [S3] |
| $\frac{d[{AlbSO}_{Periph}]}{dt}= \sigma\left[ {Alb}_{Liver} \right]-k_{AlbSO}[{AlbSO}_{Periph}]$ | Eqn. [S4] |

where ${Alb}_{Gut}$, refers to the amount of albendazole in the gut; $[{Alb}_{Liver}]$ and $[{Alb}_{Periph}]$ refer to the concentrations of albendazole in the liver and peripheral blood respectively, and $[{AlbSO}_{Periph}]$ the concentration of the metabolite albendazole sulfoxide in the peripheral blood. $k_{Abs}$ is the rate of absorption of albendazole from the gut into the bloodstream (and implicitly includes a conversion translating the absolute amount of albendazole absorbed from the gut to the corresponding concentration in the liver compartment); $sigma$ denotes the rate at which albendazole is converted to albendazole sulfoxide by the liver; $k_{Alb}$ and $k_{AlbSO}$ are the rates at which albendazole and albendazole sulfoxide, respectively, are processed and cleared in the body; $Q$ represents the rate of exchange between the liver and the peripheral blood, and for the purposes of the results presented here is set to 15 (reflecting the approximate extent of exchange anticipated per hour between the two compartments [35]). The dose of albendazole used is further multiplied by another parameter, $bioavailability$, which corresponds to the proportion of the albendazole dose received that is available for absorption.

***Model Fitting and Inferential Framework***

The above pharmacokinetic mathematical model was fitted within a Bayesian framework. Specifically, the model was fitted to each dataset individually, using an adaptive Metropolis-Hastings Markov Chain Monte Carlo (MH-MCMC) sampling algorithm developed previously [36]. Weakly informative priors were set over $k_{Abs}$ and $sigma,$ (i.e. the two parameters where further inference and association with collated metadata in the form of multiple linear regression was not carried out). For the other parameters, $bioavailability$, $k_{Alb}$, and $k_{AlbSO}$, uninformative priors were used. Prior distributions for the estimated parameters were defined as follows,

$$k_{Abs} \sim Normal(Mean= 5, Variance=7)$$

$$bioavailability \sim Normal(Mean= 0,Variance= 1)$$

$$sigma \sim Normal(Mean= 15, Variance= 7)$$

$$k_{Alb} \sim Normal(Mean= 0, Variance= 1)$$

$$k_{AlbSO} \sim Normal(Mean= 0, Variance= 1)$$

truncated at 0 so that only positive parameter values were accepted. For both albendazole and albendazole sulfoxide blood concentrations, a Poisson likelihood (reflecting the assumption that the drugs are well-mixed within each of our modelled compartments) was used, such that the model likelihood could be constructed as follows,

$$\left[ Alb \right]_{i}\sim Poisson\left( \left[ \tilde{Alb} \right]_{i} \right)$$

$$\left[ AlbSO \right]_{i}\sim Poisson\left( \left[ \tilde{AlbSO} \right]_{i} \right)$$

where $\left[ Alb \right]_{i}$ and $\left[ AlbSO \right]_{i}$ represent the empirically observed blood concentrations of albendazole and albendazole sulfoxide respectively at time-point $i$. $\left[ \tilde{Alb} \right]_{i}$ and $\left[ \tilde{AlbSO} \right]_{i}$ represent the modelled blood concentrations of albendazole and albendazole sulfoxide, respectively, at timepoint$i$. For each of the 92 time-series, a total of 50,000 iterations of the MCMC sampling algorithm were run for purposes of model fitting and parameter inference. Half of each chain’s iterations were discarded as burn-in/the adaptive phase of the sampling, leaving a total of 25,000 iterations available for inference.

***Pharmacokinetic Parameter Estimation and Multiple Linear Regression Modelling***

We next sought to associate the estimates of pharmacokinetic parameters obtained from fitting the above model, to collected metadata associated with each time-series (describing aspects of the patient population and treatment regimen received), to assess the influence of these factors on variation in albendazole and albendazole sulfoxide’s pharmacokinetics. These pharmacokinetic parameters were $k_{AlbSO}$ (related to the half-life of albendazole sulfoxide), the bioavailability of albendazole (the proportion of administered albendazole absorbed from the gut into the blood), $C_{Max}$ (the peak concentration of the drug in the blood) and $AUC$ (reflecting the total exposure to the drug after administration of the dose, calculated over a time-period of 50 hours).

For each time-series, we calculated the median value of $bioavailability$ and $k_{AlbSO}$directly from the MCMC chains generated during model fitting. However, we were unable to calculate $C_{Max}$ and $AUC$ directly from the fitted model output as studies differed substantially in the size of the dose administered (which would directly affect estimates of these two quantities). Therefore, we used the median estimates of each model parameter from the model fitting process described above, and for each time-series, simulated a hypothetical pharmacokinetic curve assuming a standardised dose of 400mg. From this hypothetical curve, standardised to have the same dose as all other time-series, we then calculated $C_{Max}$ and $AUC$; we subsequently refer to these quantities as $C_{Max400}$ and ${AUC}_{400}.$

Using a multiple linear regression-based approach, we then associated each of these pharmacokinetic parameters with the suite of collated individual/group metadata described in further detail in **Supplementary Table S1**. When examining the impact of different specific infectious diseases, we replaced the infection status variable with binary indicators for onchocerciasis (caused by infection with *Onchocerca volvulus*), echinococcosis (caused by infection with *Echinococcus granulosus* or *E. multilocularis*) and neurocysticercosis (caused by infection with *Taenia solium* (where 1 indicates that individuals or group of individuals has that infection and 0 indicates an absence of the particular infection).

**Sensitivity Analysis**

As a sensitivity analysis, we repeated the multivariate linear regression analysis described above but this time by controlling for the dose of albendazole received per kilogram of body weight (available only for a subset of the time-series due to a lack of complete information about participants’ weight), rather than the raw amount (in mg, not standardised by body weight) given to an individual (**Supplementary S2 Table**).

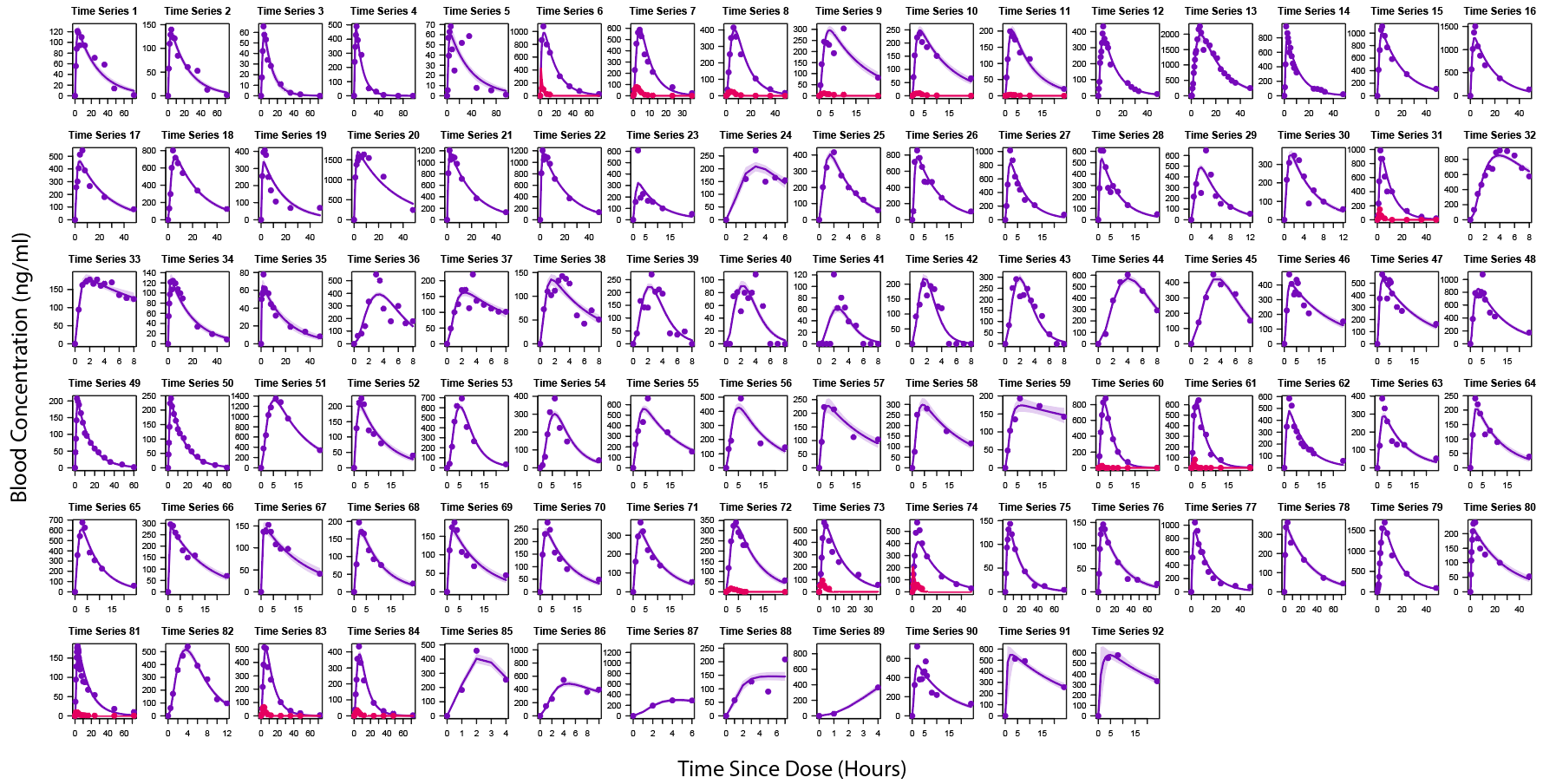

**Supplementary Figure S1: Results of model fitting and calibration to data collated through the systematic review.** The systematic review identified a total of 92 time-series containing information on the concentration of albendazole and/or albendazole sulfoxide in the blood following treatment with a single oral dose. The pharmacokinetic model of Eqns. [S1–S4] was fitted to these data individually using a Bayesian MCMC-based framework. This fitting was carried out in order to estimate the various pharmacokinetic parameters governing the model. For the results presented above, points represent empirical data and the lines represent model output, with the results for albendazole in pink and those for albendazole sulfoxide in purple. Pale shaded areas represent the 95% Bayesian Credible Interval.

**Supplementary Table S2: Multiple linear regression results relating pharmacokinetic properties to study characteristics when controlling for dosage per kilogram of body weight instead of raw dosage amount in milligrams.** Inferred pharmacokinetic parameters, specifically albendazole bioavailability, albendazole sulfoxide half-life, *C_Max_* and *AUC* were regressed onto various characteristics of the study populations controlling for sex, feeding status, age, dose per kilogram of body weight, presence of other infections (including breakdown by whether or not that infection is onchocerciasis, echinococcosis or neurocysticercosis) and co-administration of other drugs.

|  | **Bioavailability** | **AlbSO Half-Life** | ***AUC* (For Standardised 400mg Dose)** | ***C_Max_* (For Standardised 400mg Dose)** |
| --- | --- | --- | --- | --- |
| **Sex**  **(Male = Ref)** | p=0.27 | p=0.67 | p=0.51 | p=0.22 |
| **Fatty Meal** | p=0.003 | p=0.93 | p=0.005 | p<0.001 |
| **Age Group**  **(Adults = Ref)** | p=0.43 | p=0.008 | p=0.22 | p=0.52 |
| **Dose (Per kg body weight)** | p=0.09 | p=0.20 | p=0.19 | p=0.11 |
| **Parasitic Infection**  **(Ref = None)** | p=0.73 | p=0.06 | p=0.51 | p=0.95 |
| **🡺 Onchocerciasis** | p=0.31 | p=0.42 | p=0.94 | p=0.28 |
| **🡺 Echinococcosis** | p=0.006 | p=0.37 | p<0.001 | p<0.001 |
| **🡺 Neurocysticercosis** | p=0.96 | p=0.07 | p=0.10 | p=0.23 |
| **Co-Administered Drugs**  **(Ref = None)** | p=0.43 | p=0.97 | p=0.56 | p=0.43 |
